## Supplemental information for "Mosquito surveillance on U.S military installations as part of a Japanese encephalitis virus detection program: 2016 to 2021"

### Supporting Information

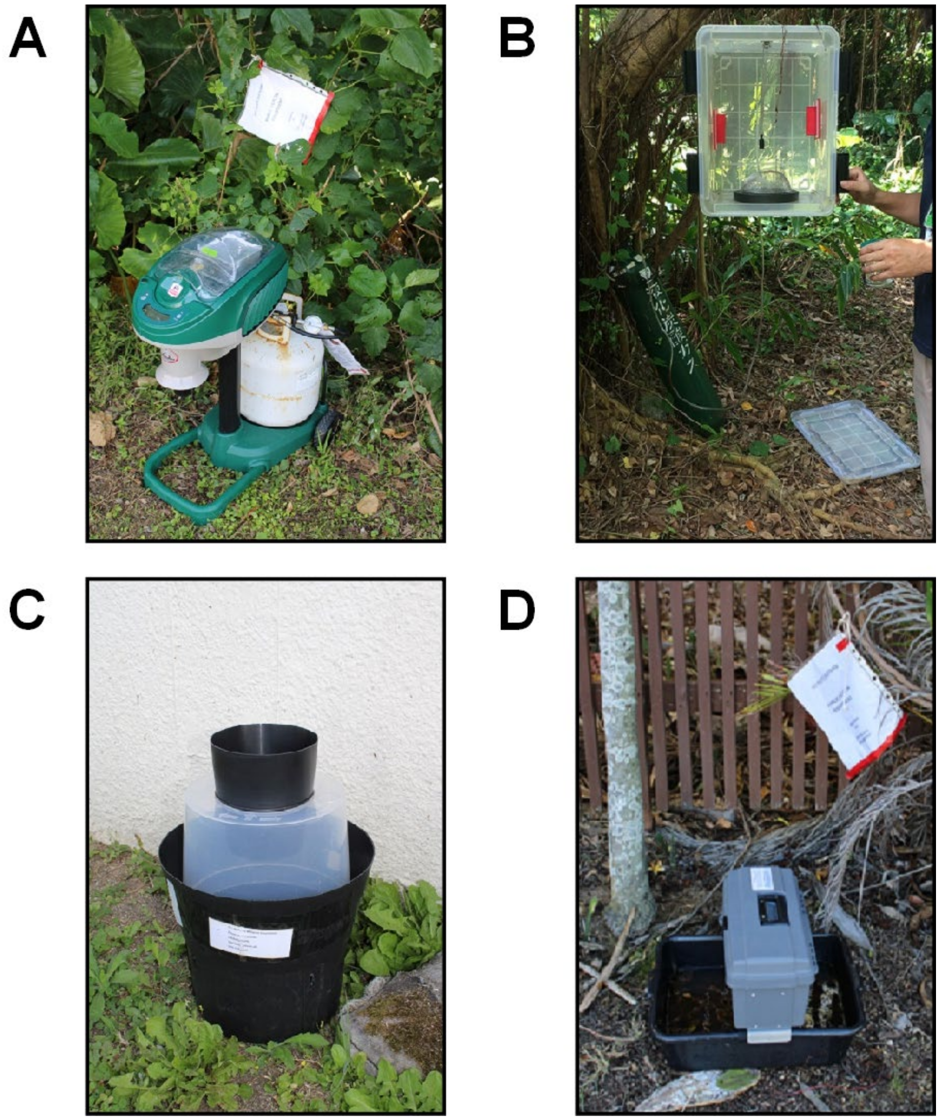


**S1 Fig.** Traps used in this study. A – Mosquito Magnet®; B – Passive Box Trap; C – Biogents Gravid Autocidal Trap; D – Reiter/Cummings Gravid Trap

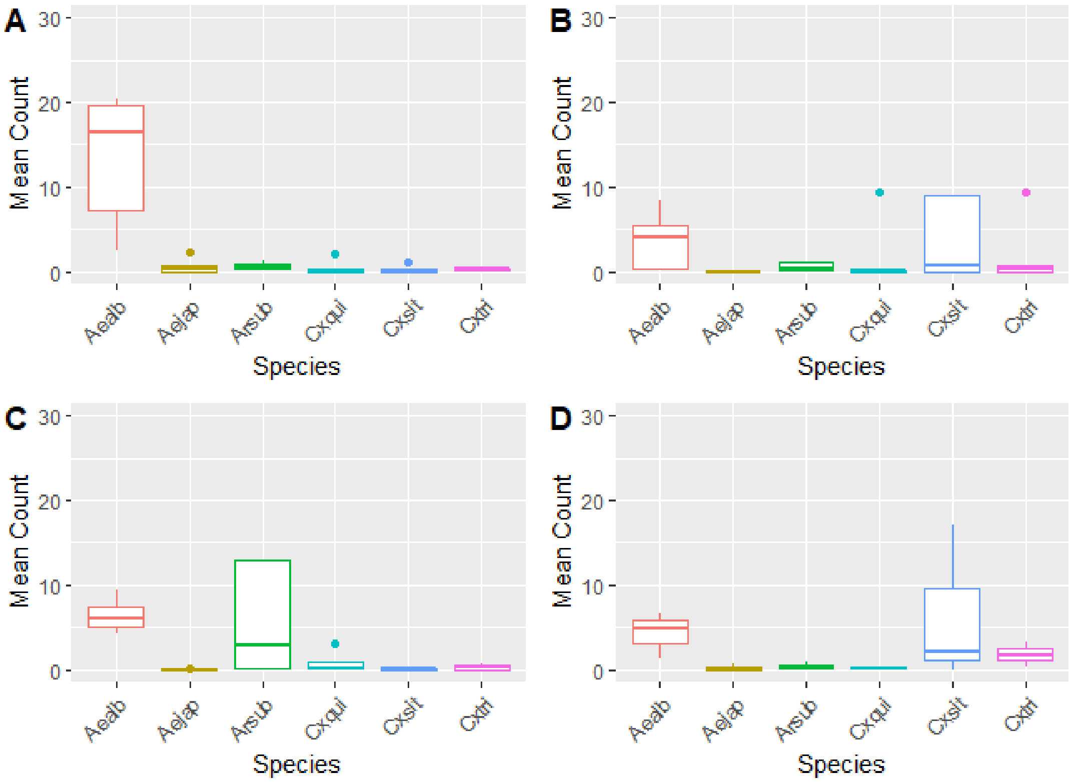


**A-** Site 1, **B**- Site 2, **C**- Site 3, **D**- Site 4

Aealb = *Aedes albopictus*; Aejap = *Aedes japonicus*; Arsub = *Armigeres subalbatus*; Cxqui = *Culex quinquefasciatus*; Cxsit = *Culex sitiens*; Cxtri = *Culex tritaeniyorhynchus*

**S2 Fig.** Mean (± SEM) count of female mosquitoes per trapping event, by species and site, 2016 – 2021.

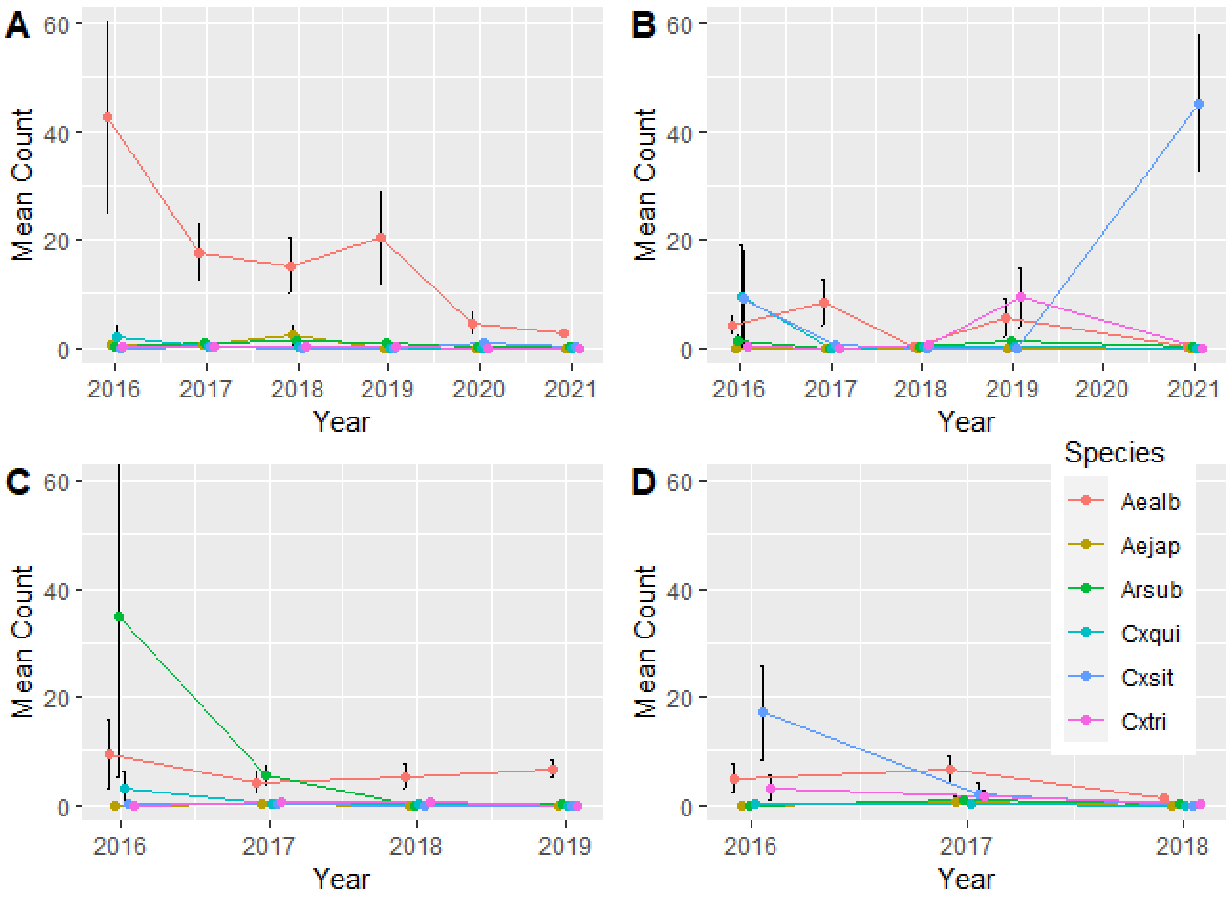


**A**- Site 1, **B**- Site 2, **C**- Site 3, **D**- Site 4

Aealb = *Aedes albopictus*; Aejap = *Aedes japonicus*; Arsub = *Armigeres subalbatus*; Cxqui = *Culex quinquefasciatus*; Cxsit = *Culex sitiens*; Cxtri = *Culex tritaeniyorhynchus*

**S3 Fig.** Change in population relative abundance over time, by species. Note: Only PBT data was used since PBT was consistently used each year of the study, at all study locations.

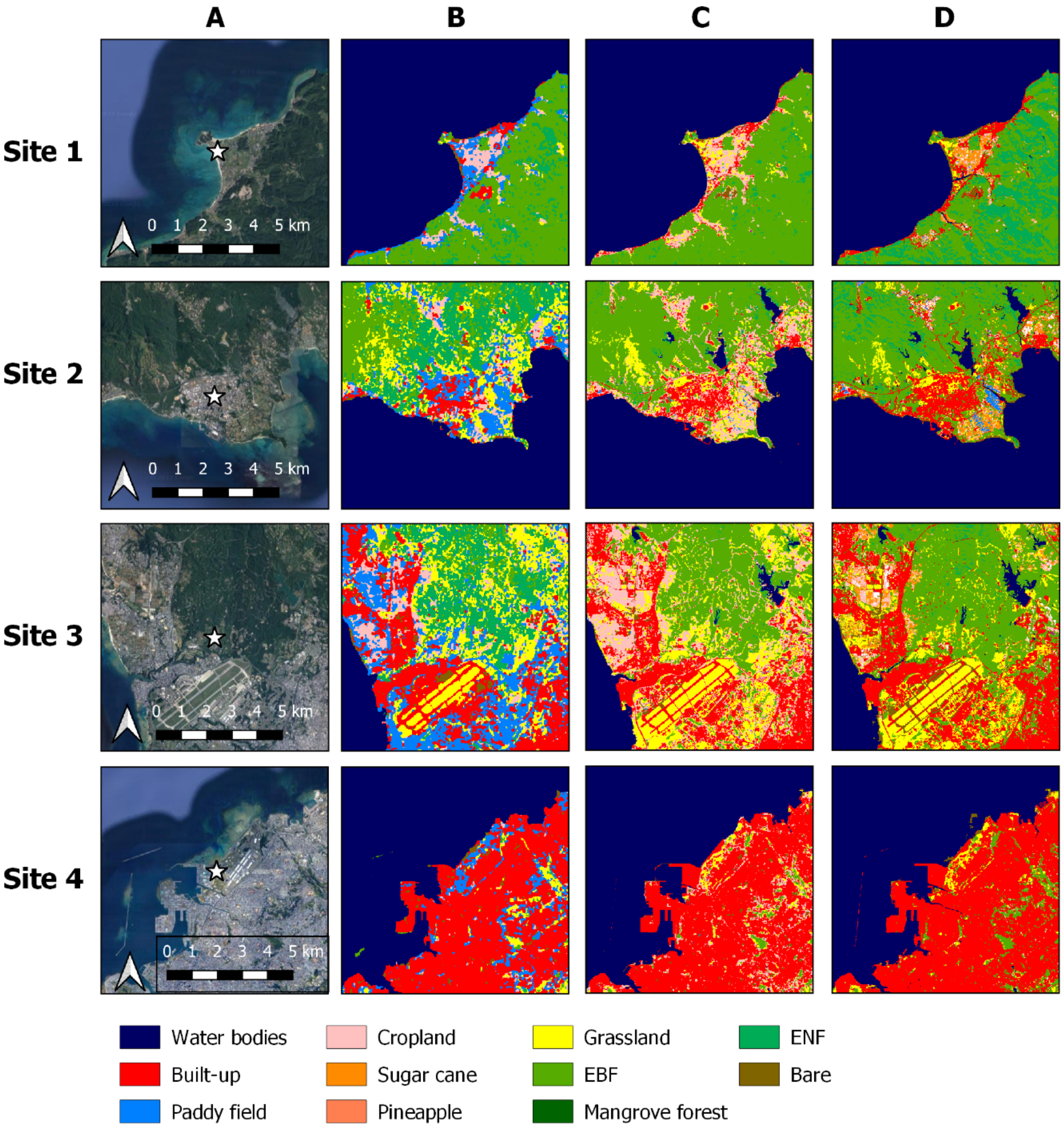


A = Satellite imagery showing trap location (☆). (Source: earth.google.com) B = Land use around 2015. C = Land use around 2019. D = Land use around 2020 (latest version). (Source: ALOS – Advanced Land Observing Satellite, Research and Application Project; https://www.eorc.jaxa.jp/ALOS/en/dataset/lulc_e.htm)

**S4 Fig.** Changes in land use at each trapping location over time.
